# Staffing levels and hospital mortality in England: a national panel study using routinely collected data

**DOI:** 10.1101/2021.12.08.21267407

**Authors:** Bruna Rubbo, Christina Saville, Chiara Dall’Ora, Lesley Turner, Jeremy Jones, Jane Ball, David Culliford, Peter Griffiths

## Abstract

**Background:** Most studies investigating the association between hospital staff levels and mortality have focused on single professional groups, in particular nursing. However, single staff group studies might overestimate effects or neglect important contributions to patient safety from other staff groups. We aimed to examine the association between multiple clinical staff levels and case-mix adjusted patient mortality in English hospitals.

**Methods and Findings:** This retrospective observational study used routinely available data from all 138 National Health Service hospital trusts that provided general acute adult services in England between 2015 and 2019. Standardised mortality rates were derived from the Summary Hospital level Mortality Indicator dataset. Estimates for the effect of clinical staffing from the single staff models were generally higher than estimates from models with multiple staff groups. Using a multilevel negative binomial random effects model, hospitals with higher levels of medical and allied healthcare professional (AHP) staff had significantly lower mortality rates (1.04, 95%CI 1.02 to 1.06, and 1.04, 95%CI 1.02 to 1.06, respectively), while those with higher support staff had higher mortality rates (0.85, 95%CI 0.79 to 0.91 for nurse support, and 1.00, 95%CI 0.99 to 1.00 for AHP support), after adjusting for multiple staff groups and hospital characteristics. Estimates of staffing levels on mortality were higher in magnitude between- than within-hospitals, which were not statistically significant in a within-between random effects model.

**Conclusions:** We showed the importance of considering multiple staff groups simultaneously when examining the association between hospital mortality and clinical staffing levels. Despite not being included in previous workforce studies, AHP and AHP support levels have a significant impact on hospital mortality. As the main variation was seen between-as opposed to within-hospitals, structural recruitment and retention difficulties coupled with financial constraints could contribute to the effect of staffing levels on hospital mortality.

## Introduction

Cost constraints and workforce shortages put pressure on health systems to find efficient staffing models to meet rising demand for care. In some countries, such as England, this has led to an increased reliance on support workforce to provide nursing care. Whilst reducing the skill mix may create short-term savings, multiple observational studies provide evidence of a risk of higher in-hospital mortality rates when registered nurse staffing levels are lowered and skill mix is diluted (1-7).

Studies focussing on the impact of clinical staff other than nurses on hospital mortality rates are relatively scarce. While some studies on medical staffing levels exist, indicating worse outcomes with lower medical staffing levels (3), most focus on differing models of rostering or deployment e.g. intensive care unit staffing models (8), physician caseload levels (9-11), or the ‘weekend effect’, whereby worse outcomes for patients admitted over weekends have been associated with having fewer experienced physicians rostered over weekend (12). To our knowledge, no recent study has examined the potential effect of allied healthcare professionals (AHP) staffing levels on clinical outcomes and only a few focused on in-hospital pharmacists (13, 14).

A small number of studies assessed the simultaneous impact of multiple staff groups on hospital mortality, even though in-hospital patient care is delivered by multidisciplinary staff and no single staff group is solely responsible for patient outcomes. Of the studies that did consider multiple staff groups, some found partial attenuation or complete absence of the effect of single staff levels on mortality when adjusted for other professionally qualified staff, indicating that uni-professional staffing estimates are potentially biased by confounding (3, 15).

In the face of enduring staffing shortages in some professional groups it is important to understand the likely consequences on patient care and to identify priorities. A focus on single professional groups risks unintended consequences through neglect of other important contributions to patient safety and might lead to biased estimates. We therefore aimed to examine the associations between staffing levels of multiple staff groups, and case-mix adjusted patient mortality in hospitals.

## Methods

### Study design

This was a retrospective observational study using routinely available data on clinical healthcare staffing and hospital mortality.

### Study setting

We included all 138 National Health Service (NHS) hospital trusts providing general acute adult inpatient services in England between April 2015 and March 2019. Hospital trusts are defined as organisational units within the NHS that service a defined geographical area or that provide a specialised function. One trust can therefore encompass several hospitals.

### Data sources and linkage

We linked four data sources that provide trust-level datasets: a) NHS workforce, which contains detailed information on trust staffing; b) bed availability and occupancy, which contains data on trust-level available and occupied beds; c) Estates Returns Information Collection (ERIC), which contains data on trust organisation and structure; and d) Summary Hospital-level Mortality Indicator (SHMI), which contains data on observed and expected deaths (S1 Table). These datasets are openly available and accessible on the NHS Digital platform, along with the data dictionary (16, 17). We linked the datasets using the unique hospital trust ID (*i*.*e*. “Org code”).

### Study outcome

The main study outcome was all-cause mortality, with standardised mortality rates derived from observed and expected deaths in the SHMI dataset (18). This is calculated and reported for all trusts providing acute adult services in NHS England (19). Specialist trusts that do not provide general acute care do not report SHMI and, therefore, were not included in the study.

The SHMI includes all in-hospital deaths and those that occurred within 30-days of discharge from patients admitted to non-specialist acute trusts. SHMI data are derived from the Hospital Episode Statistics (HES) at the level of provider spells (*i*.*e*. total continuous stay of a patient using a hospital bed at an NHS organisation under the care of one or more consultants, or nursing episode or midwife episode), and the HES-Office of National Statistics (ONS) linked mortality data (S1 Table) (20). The latter captures deaths that occur outside of hospital. Data used to calculate the SHMI are submitted by each hospital trust.

SHMI expected deaths is calculated based on individual patient characteristics that can affect the risk of mortality, including the patient’s condition for hospitalisation, underlying conditions (Charlson Comorbidity Index) (21), age, gender, method of admission to hospital, and year of discharge. Logistic regression models estimate the risk for each provider spell, with binary variable as outcome (*i*.*e*. died or survived). The SHMI model performs well with early validation accounting for 81% of between-hospital variations with a c-statistic (area under the receiver operator curve) of 0.9 (19). The models are based on the preceding three years’ data, with last year data used to calculate the SHMI (22). As SHMI data are published monthly, we used the dataset that contains mortality data from April to March the following year to report on annual hospital mortality level for each hospital trust included in this study.

### Study variables

We obtained hospital staffing data by linking the medical and dental workforce dataset with the non-medical workforce dataset. Bed occupancy data are published quarterly as averages, with no estimate of monthly variation. For wards open overnight, occupied bed is defined as when it is occupied at midnight on the day in question; for day-only wards, occupied bed is a bed where at least one day case has taken place during the day. Trust teaching status was derived from the ERIC dataset, which contains variables on trust profile.

Staffing variables are published monthly. We used full-time equivalent (FTE) values to calculate the annual average available staff at each hospital trust from April to March the following year, to align with the SHMI data. FTE data are based on the proportion of time each staff are expected to work in a week, which would correspond to an FTE of 1 (*e*.*g*. 48 hours for doctors, 37.5h for nurses). Overtime and out-of-hour work are not recorded in these datasets.

Clinical staff are classified according to occupation codes. We grouped staffing variables by frequency and occupation as listed by NHS England, merging groups that had mean below 3 with others of a similar occupation in order to create 8 groups (*i*.*e*. medical, surgical, other medical specialties, nurses, support to nurses, AHP, support to AHP, and scientific, therapeutic, and technical (ST&T) staff) (see S1 Appendix for details).

We used the number of general acute occupied beds to calculate the bed-per-staff ratio. Beds assigned to maternity, mental illness, and learning disabilities services were excluded; however, general acute beds represented 96.2% of the total beds available across all hospital trusts included in the study. We calculated the average number of occupied beds overnight or day-only per trust per year, and then divided occupied beds per each staff group to obtain the average bed-per-staff levels for each hospital trust per year.

Teaching affiliation was coded as yes or no according to the trust type recorded in the ERIC dataset. Trust size was calculated based on the number of available general acute beds in each trust. There was a median of 744.2 available general acute beds per trust (interquartile range (IQR) 535.7 to 1009.4, range 143 to 2704). Hospital trusts in the upper tertile (*i*.*e*. the upper third of trusts, ranked by size) were classified as large, while those in the lower tertile were deemed small trusts, with the remaining classified as medium-sized hospital trusts.

Observations with missing data were removed from the analyses; however, hospital trusts that reported at least one year of complete data were included. There were 540 observations in the linked dataset, of which 519 (96%) contained data on all variables and were therefore included in our analyses. There were 2 hospital trusts that did not report on medical staffing in 2016, with 6 trusts not reporting these variables from 2017 onwards. For other clinical staff, only 1 trust did not report staff numbers in 2016 and 2017, and two trusts in 2018, with no missing data in 2019. One trust did not report the number of occupied general acute beds in 2018. The number of hospital trusts that reported on complete data varied, with 137 included in 2016 and 2017, 135 in 2018, and 130 in 2019.

### Statistical analysis

We initially explored the data using descriptive statistics. Continuous variables were described as mean and standard deviation (SD) or median and IQR, depending on the distribution of each variable. Categorical variables were described as frequencies and proportions. Annual mortality rates were calculated by dividing the number of observed deaths by the number of patient spells, across all hospital trusts each year. To explore the relationship between staffing levels and hospital mortality, statistical modelling was conducted at the hospital trust-level using multiple regression models on four years of data, with standardised mortality rates regressed onto hospital level staffing levels (expressed as the number of occupied beds-per-staff). We included expected deaths as an offset, as the number of expected deaths is the number of times the event could have happened (*i*.*e*. the exposure variable).

Multilevel models were adjusted for trust size and affiliation as a teaching hospital, with trust included as a random effect to adjust for clustering. We considered a range of potential models, with model selection based on minimising the Akaike information criterion (AIC), the Bayesian information criterion (BIC) and the likelihood ratios (see S2 Table for alternative frameworks considered). We report exponentiated coefficients (rate ratios) with 95% confidence intervals (CI) for all estimates obtained from the models.

The best performing model (referred to as main model) was a negative binomial random effects model that included all 8 clinical staff groups and hospital characteristics (*i*.*e*. trust size and teaching status). To explore variations between and within hospital trusts, we also constructed a within-between random effects model (WBRE), with trusts as random effect, and staffing and hospital characteristics as fixed effects. These models can differentiate between effects arising from staffing differences between hospital trusts and those that are associated with annual changes of staffing levels within each hospital (23, 24).

We assessed potential collinearity between co-variates using correlation plots and Spearman’s correlation coefficient, and the overall multicollinearity for all predictors in the models using generalised variance inflation factors (GVIF). All models had a GVIF<10. As a sensitivity analysis, we reran the models after excluding any variables that had a GVIF above 5 (25).

We performed data linkage, cleaning, coding, and analyses in R statistical software, version 4.0.2 (R Core Team, R Foundation for Statistical Computing, Vienna, Austria), using the lme4 and glmer packages (v1.1-27.1, Bates et al, 2015), and the plm package (v2.4-3, Croissant et al, 2021) (26).

## Results

### Descriptive statistics

Our final sample consisted of 519 observations of staffing, bed occupancy and mortality variables for the 4-year study period, clustered within 138 hospital trusts. The median number of acute general occupied beds per day was 674.0 (IQR 486.1 to 911.6). The mean annual observed deaths ± SD was 2229 ± 992 (range 525 to 7468), while the expected deaths were 2228 ± 987.2 (range 665 to 7591). Annual mortality rates across all hospital trusts were 3.44% for 2016, 3.46% for 2017, 3.23% for 2018, and 3.31% for 2019. Forty-one (29.7%) hospital trusts were classified as teaching trusts.

Table 1 shows the distribution of staffing variables by their median, IQR, mean, and SD across all hospital trusts. There was considerable variation in staffing levels, with nurse staff levels showing least relative variation (SD 18.0% of the mean) and AHP support most (SD 44.4% of the mean). By contrast, variations within trusts as a percentage of the mean were relatively low, with the within trust variation ranging from 4.9% for nurse staffing to 10.9% for AHP support.

**Table 1.**
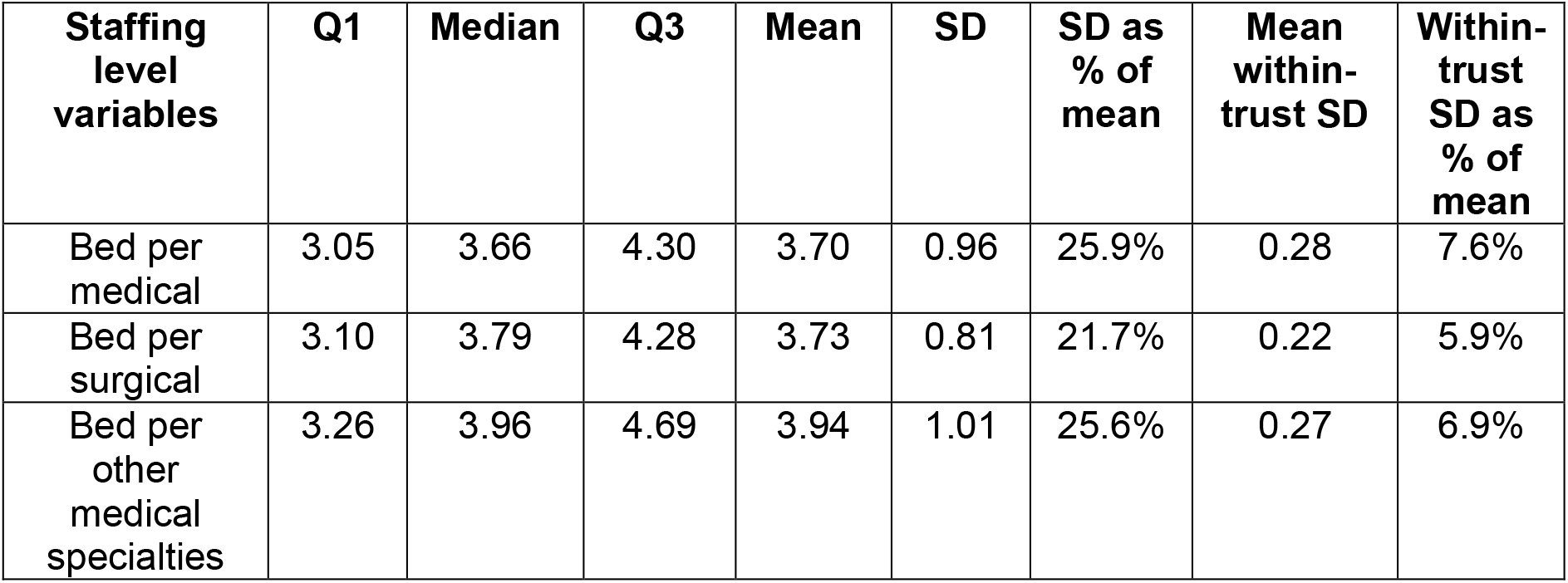
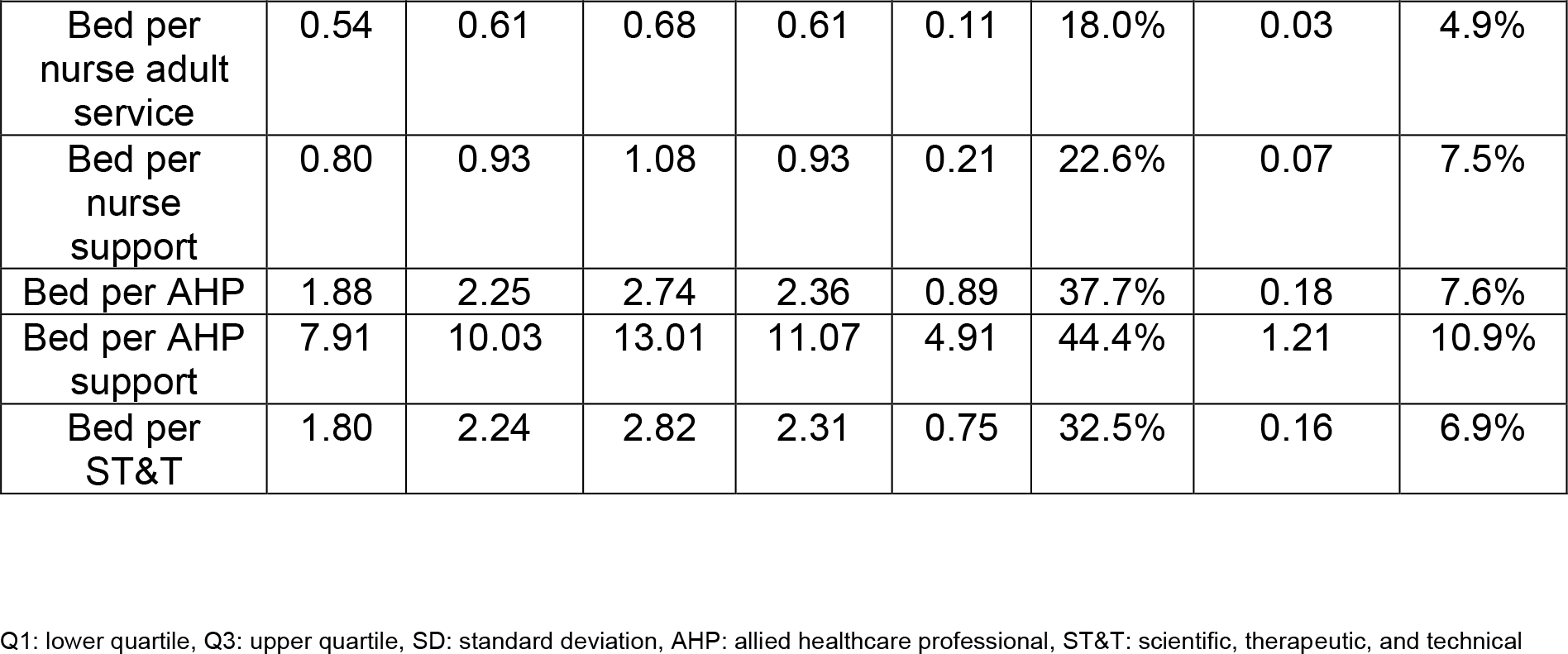
Frequency of occupied bed-per-staff variables across 138 hospital trusts

| Staffing level variables | Q1 | Median | Q3 | Mean | SD | SD as % of mean | Mean within-trust SD | Within-trust SD as % of mean |
| --- | --- | --- | --- | --- | --- | --- | --- | --- |
| Bed per medical | 3.05 | 3.66 | 4.30 | 3.70 | 0.96 | 25.9% | 0.28 | 7.6% |
| Bed per surgical | 3.10 | 3.79 | 4.28 | 3.73 | 0.81 | 21.7% | 0.22 | 5.9% |
| Bed per other medical specialties | 3.26 | 3.96 | 4.69 | 3.94 | 1.01 | 25.6% | 0.27 | 6.9% |
| Bed per nurse adult service | 0.54 | 0.61 | 0.68 | 0.61 | 0.11 | 18.0% | 0.03 | 4.9% |
| Bed per nurse support | 0.80 | 0.93 | 1.08 | 0.93 | 0.21 | 22.6% | 0.07 | 7.5% |
| Bed per AHP | 1.88 | 2.25 | 2.74 | 2.36 | 0.89 | 37.7% | 0.18 | 7.6% |
| Bed per AHP support | 7.91 | 10.03 | 13.01 | 11.07 | 4.91 | 44.4% | 1.21 | 10.9% |
| Bed per ST&T | 1.80 | 2.24 | 2.82 | 2.31 | 0.75 | 32.5% | 0.16 | 6.9% |

Q1: lower quartile, Q3: upper quartile, SD: standard deviation, AHP: allied healthcare professional, ST&T: scientific, therapeutic, and technical

Nurse staffing levels were strongly correlated with staffing levels in all medical staff groups (rho>0.71). Nurse support staff levels were correlated to RN (rho=0.30), as were AHP and AHP support (rho=0.47).

### Staffing levels and hospital mortality

Table 2 shows results for the negative binomial random effect models, adjusted for hospital characteristics. In the single staff group models, hospital trusts that had lower staffing levels (*i*.*e*. more occupied beds per FTE staff) were associated with higher (standardised) mortality rates for all professionally qualified staff groups. The opposite effect was observed for nursing support and AHP support staff, where hospital trusts with lower staffing levels (*i*.*e*. more occupied beds per FTE staff) had lower mortality.

**Table 2.** Exponentiated estimates for hospital mortality from the multilevel negative binomial random effect models, adjusted for hospital characteristics (*n* = 138)

| Predictors | Estimates for single staff models | 95% CI | Estimates for multiple staff model | 95% CI |
| --- | --- | --- | --- | --- |
| Bed per medical | 1.05*** | 1.04 to 1.07 | 1.04*** | 1.02 to 1.06 |
| Bed per surgical | 1.04*** | 1.02 to 1.06 | 0.98 | 0.96 to 1.01 |
| Bed per other medical specialties | 1.05*** | 1.03 to 1.06 | 1.03* | 1.00 to 1.06 |
| Bed per nurse adult service | 1.33*** | 1.15 to 1.54 | 1.07 | 0.88 to 1.31 |
| Bed per nurse support | 0.92* | 0.86 to 0.98 | 0.85*** | 0.79 to 0.91 |
| Bed per AHP | 1.02* | 1.00 to 1.04 | 1.04*** | 1.02 to 1.06 |
| Bed per AHP support | 0.99* | 0.99 to 0.99 | 1.00** | 0.99 to 1.00 |
| Bed per ST&T | 1.02** | 1.00 to 1.04 | 0.99 | 0.98 to 1.05 |
| Teaching (ref not teaching) | 0.96* | 0.92 to 0.99 | 1.01 | 0.98 to 1.05 |
| Medium trust (ref small trust) | 1.01 | 0.97 to 1.05 | 1.01 | 0.98 to 1.04 |
| Large trust (ref small trust) | 1.00 | 0.96 to 1.04 | 1.01 | 0.98 to 1.05 |

CI: confidence interval, AHP: allied healthcare professional, ST&T: scientific, therapeutic, and technical, \* $p < 0.05$ , \*\* $p < 0.01$ , \*\*\* $p < 0.001$

In the main multivariable model including all staff groups, associations between medical (1.04, 95%CI 1.02 to 1.06), other medical specialties (1.03, 95%CI 1.00 to 1.06), AHP (1.04, 95%CI 1.02 to 1.06), nurse support (0.85, 95%CI 0.79 to 0.91), and AHP support (1.00, 95%CI 0.99 to 1.00) FTE staffing levels and hospital mortality remained statistically significant, with no change in direction of the effect compared to the single staff models (Table 2). Higher mortality rates were observed in hospital trusts with lower levels of medical and AHP staff. In contrast, hospital trusts with lower support staff levels per occupied bed (*i*.*e*. nursing support and AHP support) had lower mortality rates. The association with RN staffing was attenuated and no longer statistically significant (1.07, 95%CI 0.88 to 1.31), although the observed effect remained relatively large compared to other associations (S1 Fig). Similarly, the association between hospital mortality and ST&T staff, observed in the single staff groups models, were no longer statistically significant in the model adjusted for all staff groups.

Although we noted correlation between staffing variables, our GVIF for all variables was <10 which is generally accepted as indicative of no evidence of multicollinearity (27). The GVIF for beds per other medical specialties was 6.1 (S3 Table), and therefore over our threshold of 5, so we ran the negative binomial random effects model omitting this variable as a sensitivity analysis and found similar results (not shown; available from authors upon request).

### Within-between hospital trust variability

Between-hospital trust effects from the WBRE model were largely the same as those obtained from the main model (*i*.*e*. between only). For example, the between-estimate from WBRE for the FTE medical group was 1.04 (95%CI 1.02 to 1.07), similar to the estimates obtained from the main model (1.04, 95%CI 1.02 to 1.06). However, even for staff groups where between-hospital trusts effects were significant, the within-hospital effects were small and not statistically significant. For example, the within-hospital estimate from the WBRE model for the medical group was 0.99 (95%CI 0.97 to 1.01) (Fig 1). Despite potentially providing additional information by decomposing the variability into between- and within-estimates, the WBRE model did not improve fit compared to the main model (S2 Table).

**Figure 1.**
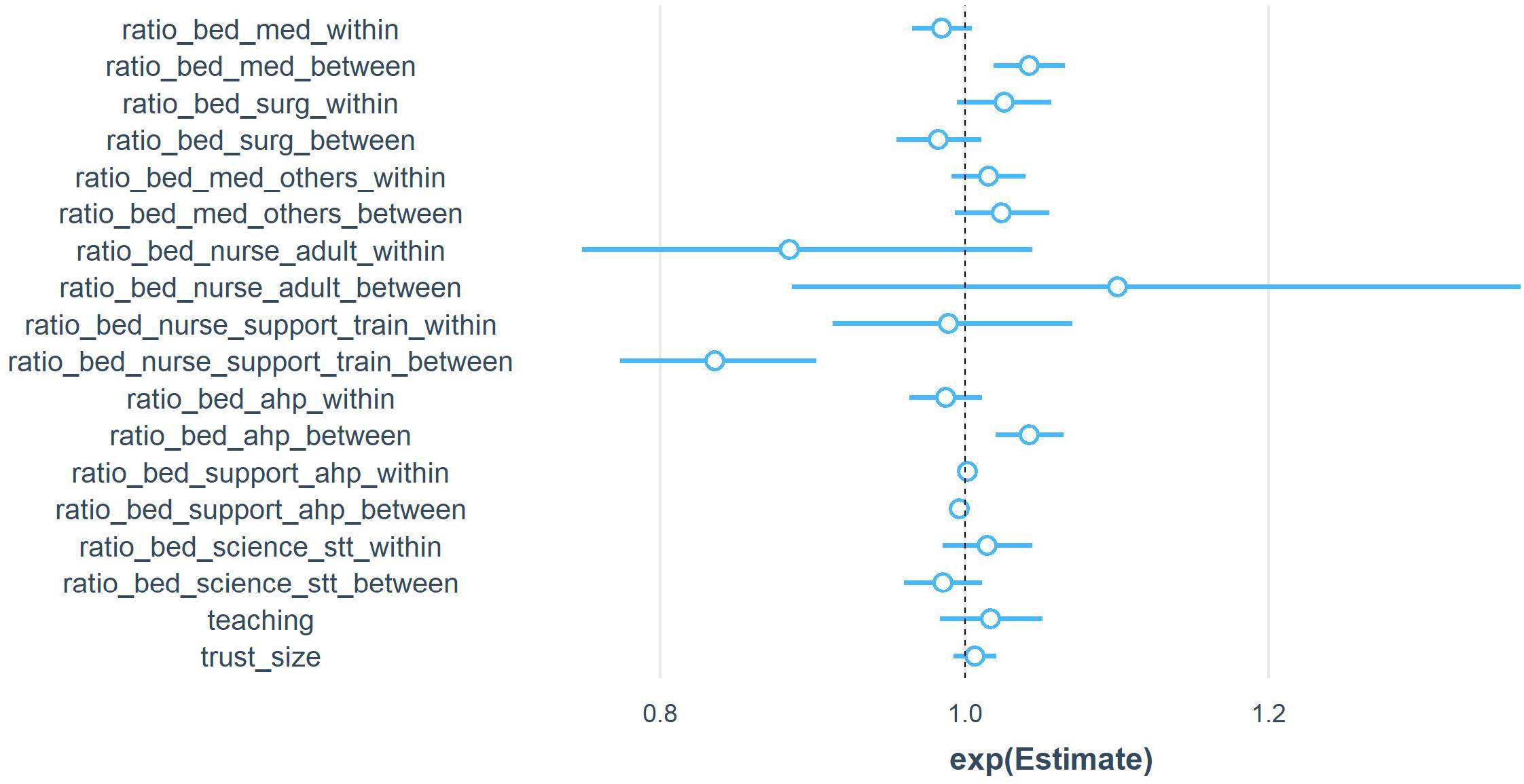
Hospital mortality exponentiated estimates obtained from the within-between random effect model (WBRE).

## Discussion

After adjustment for hospital characteristics and including multiple clinical staff groups in our models, we found that higher medical and AHP staffing levels were associated with lower mortality rates in acute hospital trusts in England during the period of 2015 to 2019. In contrast, hospitals with higher support staff per occupied bed had higher mortality rates.

Our study highlights the importance of simultaneously adjusting for multiple clinical staff groups when investigating associations between staffing levels and mortality at the hospital-level. In the single staff group models, significant effects on mortality rates were seen for all staffing groups. However, when adjusted for multiple groups, effects for medical, other medical specialties, AHP, nurse support, and AHP support staffing levels remained statistically significant, but levels of surgical doctors, RN, and ST&T staff were no longer significant.

Our findings are in line with previous research. Griffiths *et al* (15) reported that higher levels of occupied beds per RN and per doctor were associated with increased mortality in the single staff models, while the association was reversed for healthcare support workers. However, there was a substantial attenuation in nurse staffing estimates when medical staffing levels were included. Jarman *et al* (3) showed that the association between RN staffing and mortality was no longer significant when adjusting for doctors-per-bed, with higher levels of physician staffing significantly reducing hospital standardised mortality ratios.

Other studies have shown a reduction in hospital mortality when higher numbers of doctors and nurses per bed were observed (12, 15, 28-33). In our study, although the largest estimate of effect on patient mortality was for nurse staffing, this was not statistically significant in the main model, which was adjusted for all clinical staff groups. Studies have reported that much of the variation in nurse staffing occurs between wards within hospitals (34, 35) and Keogh noted that hospital nurse staffing levels often bore little relationship to staffing available to be deployed on wards in NHS hospitals (36), therefore hospital nurse staffing per bed may be a poor indicator of the staffing levels experienced by inpatients. Griffiths *et al* (15) found that when medical staffing levels were included in models, significant nurse staffing effects were only observed in models using ward-based staffing ratios as opposed to hospital-level staff-per-bed ratios. Longitudinal patient level studies of exposure to variation in nurse staffing confirm that there is an adverse effect of low nurse staffing when measured at this level (37-39).

In our study, high levels of nurse support staff were associated with higher mortality rates. Bond *et al* (28) showed that higher staffing levels of medical residents, RN, pharmacists, and medical technologists resulted in lower mortality rates, while nurse support and administrators had the opposite effect. The reasons for the reverse effect of support staff on mortality are unclear; however, they could include reduction of skill mix by increasing the proportion of support staff in relation to professionally qualified staff, and increased workload of professionally qualified staff due to additional supervisory tasks when support staff levels are higher (40).

To our knowledge, this was the first study to include AHP and AHP support staff levels in analysis. Higher levels of AHP staff had a protective effect at the hospital trust level while more AHP support staff were detrimental to patient mortality, a finding that mirrors those obtained for nursing support staff. While causality cannot be assumed for any of these results, there is a potential that previous findings that have emphasised a link between nurse staffing and patient safety could divert attention from the role that other staff groups may play in delivering quality and preventing avoidable deaths.

We found that variations between hospital trusts in the 4-year study period were higher in magnitude than within-hospital trusts estimates, which were not statistically significant for any of the clinical staff groups at the hospital level. Structural recruitment and retention difficulties, and financial constraints could be main contributors to inter-hospital trust variations. There was relatively little year on year variation in staffing between trusts. Reduced variability of staffing levels within hospitals might be due to the limited number of funded posts in each organisation and could reflect historic staffing levels. Bjerregaard *et al* (33), using a WBRE model, found staffing levels estimates were significant between wards, while within-ward estimates had no significant effect on hospital mortality.

### Strengths and limitations

This was a national study using 4 years of routinely available data. We considered the effects of staffing by professionally qualified and support workers on in-hospital and 30-day mortality rates. The inclusion of numerous clinical staffing groups simultaneously in the models allowed us to estimate the effect of each staffing level on hospital mortality while considering all other clinical staff levels and hospital trust characteristics.

We used a variety of models and selected the one that performed best on our data. The use of different methodological approaches highlighted how different assumptions about the distribution of staffing variables and the nature of staffing between and within hospitals can impact model performance and findings derived from such models. Most studies investigating clinical staffing levels and in-hospital mortality used fixed effects models, which only evaluate within-hospital effects. While these might be useful when estimating the impact of interventions on staffing levels within a hospital, they might not appropriately assess the effect of different staffing levels based on variations between hospitals in hospital-level observational studies. We showed that between-hospital effects are larger and generally dictate the relationship between clinical staffing levels and mortality. In fact, some of the estimates of within-hospital staffing levels had the opposite direction to the between-hospital estimates on hospital mortality.

However, our study was limited by the relatively small sample size, as observations were clustered on 138 hospital trusts. We therefore had to group some of the medical specialties and were unable to unpack relatively heterogenous groups such as AHP. Furthermore, we were unable to explore the effects of grade and experience for medical (*e*.*g*. junior doctors, level of consultants) and RN (*e*.*g*. senior nurses) staff.

In our study, beds that were occupied by multiple patients in a single day were counted as one single occupied bed per staff. Studies have shown that workload (*e*.*g*. number of admissions, discharges, additional tasks) in high volume hospitals can lead to higher mortality rates, even when levels of staffing are similar to those observed in low volume hospitals. Occupied beds-per-staff might not be a good measure of staffing levels, as these do not take into account temporary staff or staff absence due to sickness, maternity leave or long-term leave. The extent to which staff are deployed to deliver services such as ambulatory care or other services not delivered to inpatient beds is likely to vary (41).

Future studies should aim to capture different staffing levels responsible for delivering patient care measured at the patient or ward-level, rather than at the hospital-level.

Although SHMI provides good control for patient risk and we controlled for a number of hospital factors and multiple staff groups, the cross-sectional nature of our analysis means that findings need to be interpreted as demonstrating association but not direct evidence of causation.

## Conclusion

In conclusion, our study highlights the importance of simultaneously considering multiple staff groups when investigating the effect of clinical staffing levels and mortality at the hospital-level. We showed that the number of AHP and AHP support per occupied beds have a significant impact on patient mortality, yet these groups have largely not been included in previous workforce studies, of which the majority focused exclusively on nursing staff. We also found that hospitals with higher levels of medical staffing had lower mortality rates, while higher levels of support workers were associated with higher hospital mortality.

## Data Availability

"All data supporting this study are openly available from the University of Southampton repository at https://doi.org/10.5258/SOTON/D2055"

## Supporting information captions

Table S1. NHS digital data sources

Table S2. Comparison of goodness-of-fit for model selection

Table S3. Multicollinearity test in the main model (*i*.*e*. negative binomial random effect)

**Figure S1.**
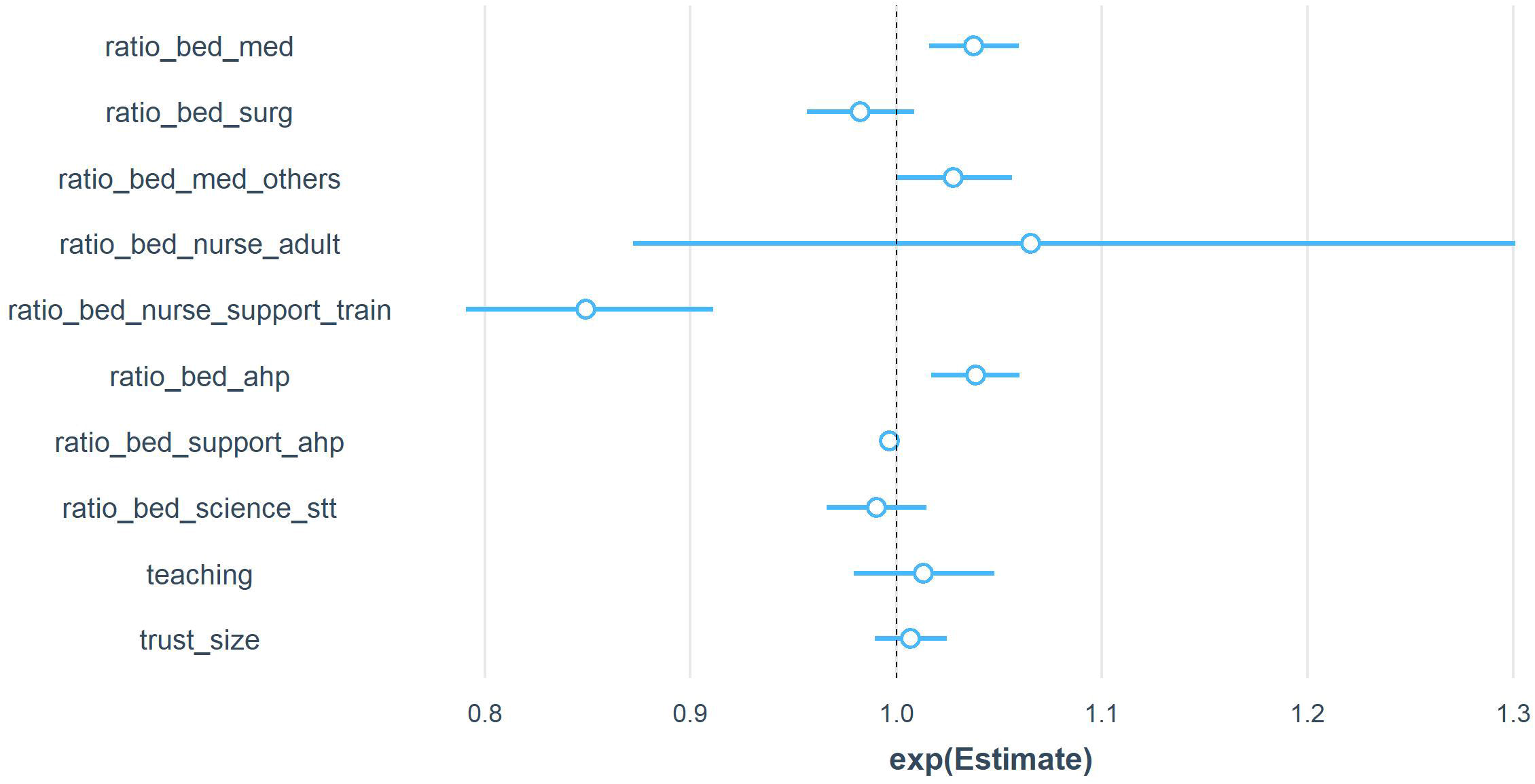
Hospital mortality estimates obtained from the main model (*i*.*e*. negative binomial random effect).

## Notes

### Competing Interest Statement

The authors have declared no competing interest.

### Funding Statement

This study is funded by the National Institute for Health Research (NIHR) [Health Services & Delivery Research (NIHR128056)]. The views expressed are those of the author(s) and not necessarily those of the NIHR or the Department of Health and Social Care.

### Author Declarations

All data used in this study are openly available and can be access through NHS Digital at https://digital.nhs.uk/data

