## Supplementary materials for "Staffing levels and hospital mortality in England: a national panel study using routinely collected data"

### Supporting information

#### Staff variable groups

General medicine specialist doctors were grouped with clinical oncology doctors to form the general medicine group, surgeons were grouped with obstetricians and gynaecologists to form the surgical group, and all other medical specialties were grouped together to form the other medical group (*i.e.* radiologists, emergency medicine, anaesthetists, and pathologists). We excluded psychiatrists and paediatricians. The nurses’ group was composed of nurses in adult services only; we did not include nurses from paediatric services. The nurse support group included support to adult and general nurses, and nurses in training. Staff in the fields of podiatry and chiropody, dietetics, occupational therapy, orthoptics and optics, physiotherapy, radiography, art, music and drama therapy, and speech and language therapy were included in the allied health professional (AHP) group. Scientific, therapeutic, and technical (ST&T) staff group was composed of multi-therapists, applied psychologist, psychological therapist, pharmacists, dentists, operating theatre staff, social services, and other ST&T staff.

#### Tables and figures

Table S1. NHS digital data sources

| **Data source** | **Datasets** | **Main variables available** |
| --- | --- | --- |
| NHS workforce data | Medical dataset | Professionally qualified clinical staff |
|  | Non-medical dataset | Professionally qualified clinical staff |
|  |  | Support to clinical staff |
| Bed occupancy data | Overnight beds dataset | Available beds |
|  |  | Occupied beds |
|  |  | % occupied beds |
|  | Day only beds datasets | Available beds |
|  |  | Occupied beds |
|  |  | % occupied beds |
| ERIC data |  | Trust profile (e.g. type) |
|  |  | Strategies and policies |
|  |  | Finance |
|  |  | Safety |
|  |  | Fire and safety |
|  |  | Business transport |
|  |  | Medical records |
|  |  | Key worker accommodation |
|  |  | Facilities management services |
|  |  | Areas |
|  |  | Function and space |
|  |  | Quality of buildings |
|  |  | Combined heat and power energy |
|  |  | Energy |
|  |  | Water services |
|  |  | Waste |
| SHMI data |  | SHMI value |
|  |  | SHMI banding |
|  |  | Number of spells |
|  |  | Observed deaths |
|  |  | Expected deaths |

NHS: National Health Service, ERIC: Estates Returns Information Collection, SHMI: Summary Hospital level Mortality Indicator

Table S2. Comparison of goodness-of-fit for model selection

| **Model** | **Correlations for trusts** | **Model degrees of freedom** | **AIC** | **BIC** | **Likelihood-ratio** |
| --- | --- | --- | --- | --- | --- |
| OLS | No | 17 | 8183.5 | 8255.8 | -4074.7 |
| Poisson | No | 16 | 10567.6 | 10635.7 | -5267.8 |
| Negative binomial | No | 17 | 6692.6 | 6764.9 | -3329.3 |
| GLS | Yes | 26 | 7517.0 | 7627.6 | -3732.5 |
| GLS exchangeable | Yes | 18 | 7867.5 | 7944.0 | -3915.7 |
| GLS Toeplitz (2) | Yes | 19 | 7823.9 | 7904.7 | -3892.9 |
| GLS AR | Yes | 18 | 7822.7 | 7899.2 | -3893.3 |
| Poisson RE | Yes | 14 | 1997.8 | 2057.3 | -984.9 |
| **Negative binomial RE** | **Yes** | **14** | **250.3** | **309.9** | **-111.2** |
| Poisson FE | Yes (dummy) | 146 | 6477.4 | 7098.2 | -3092.7 |
| Negative binomial FE | Yes (dummy) | 147 | 6180.5 | 6805.6 | -2943.3 |
| Negative binomial WBRE | Yes | 22 | 6418.3 | 6511.8 | -3187.1 |

AIC: Akaike information criterion, BIC: Bayesian information criterion, OLS: ordinary least squares, GLS: generalised least squares, AR: autoregressive, RE: random effects, FE: fixed effects, WBRE: within-between random effects, (2) tridiagonal 2-Toeplitz matrix

Table S3. Multicollinearity test in the negative binomial random effects model

| **Variable** | **GVIF** |
| --- | --- |
| Beds per medical | 3.1 |
| Beds per surgical | 3.6 |
| Beds per other medical | 6.1 |
| Beds per nurse | 3.7 |
| Beds per nurse support | 1.7 |
| Beds per AHP | 2.0 |
| Beds per AHP support | 1.4 |
| Beds per ST&T | 2.6 |
| Teaching status | 1.8 |
| Trust size | 1.1* |

GVIF: generalised variance inflation factor; AHP: allied health professionals; ST&T: scientific, therapeutic, and technical; *reported on GVIF^(1/(2*Df)) instead of GVIF as variable has 2 degrees of freedom
